# Depressive Symptoms and Negative Experiences in School: A Network Analysis

**DOI:** 10.1101/2021.03.23.21254089

**Authors:** Yi Huang, Petr Macek, Jinjin Lu

## Abstract

**BACKGROUNDS:** Negative experiences in school predict youth’s depression. However, the dynamic interactions of depressive symptoms with adolescents’ negative experiences in school remain unclear. This study aimed to applied network analysis to detect the complex relationships between early adolescents’ depressive symptoms and negative experiences in school.

**METHODS:** We adopted the data from a Chinese national survey conducted in 2018. Eight hundred ninety-seven adolescents from 10 to 15 years old were included. The measurements included an 8-item depression screener scale and a 14-item scale assessing negative experiences in school.

**RESULTS:** The centrality analysis suggested that Chinese adolescents’ core depressive symptoms were negative affections and negative cognition that “ I felt that everything I did was an effort. “ Regarding “negative experiences in the school,” the most central nodes were poor academic performance and peer relationships. The bridge-centrality results showed negative emotions in school and difficulties in peer relationships were significantly linked to depression.

**CONCLUSION:** Negative emotional experiences in school and peer relationships were the two most important factors linked to depression.

## BACKGROUND

Depression is a common major depressive disorder. As one of the most important psychiatric diagnosis in the world, the fifth edition of The Diagnostic and Statistical Manual of Mental Disorders (DSM-V) outlines several criteria for depression, including that “the individual must be experiencing five or more symptoms during the same 2-week period and at least one of the symptoms should be either (1) depressed mood or (2) loss of interest or pleasure” ^1^. In 2017, the World Health Organization reported that more than 300 million people were suffering from depression. It is worth noticing that adolescents could experience depression as well. The previous research has reported “an estimated one-year prevalence of 4–5% in mid to late adolescence” ^2^.

School experience plays a significant role in the youth’s mental health. According to the ecological system theory in developmental psychology ^3^, the individual’s environment directly influences both cognitive and affective development. For children and adolescents, the main environments are school and family. Adolescents’ experiences in school are strongly associated with their mental health, including depressive traits. About 50 years ago, an American psychologist ^4^ contemplated that negative experience in school predicts adolescents’ and children’s depression. He showed some empirical evidence supporting a strong relationship between depression and emotional problems in school, such as feeling rejected and guilty ^5^ and feeling sad and lonely ^6^. Additionally, academic performance is an indicator of depression, such as poor concentration and academic achievement ^7^. The theory is still applicable in the 21st century. For example, psychologists suggest school-aged children’s depression is associated with their concentration difficulties, peer relationships, and academic achievement in schools ^8^. Likewise, an interview study pointed out that adolescents with depression face many learning and social interaction challenges in school ^9^.

However, not all studies on adolescents’ depression and their experience in schools considered the cultural context. Most of them were conducted in the western context. Even though very few studies have investigated how academic achievement predicts adolescents’ depression in the Chinese context, conclusions were controversial due to regional differences. Hesketh et al. ^10^ noted that in urban areas of China, but not in rural areas, poor academic performance contributes to suicide ideation caused by depression. However on the other hand, a regional study in China ^11^ found that higher self-reported academic performance indicates less depression. However, the related studies that have provided comprehensive measurement of behaviors or experiences in schools have been insufficient. This research utilized the national data to gather the perspectives on Chinese early adolescents’ depressive symptoms and probe the link between depressive symptoms and their multi-negative experiences in schools. We aimed to find out the core symptoms of depression and the core negative experiences in schools. Next, we explored negative experiences that can potentially link to depression.

## METHODS

### Participants

This study utilized the data of 897 adolescents from a Chinese public database named Chinese Family Panel Studies (CFPS). CFPS project has initiated since 2010, which is designed to collect individual-, family- and community-level data every two years to provide information on Chinese development of economic activities, family relationships, educations, and citizen’s health (including physical health and mental health). This project adopted the multi-stage stratified sampling method, which includes 25 provinces in China covering over 95% population ^12^. The data in the current study were collected in 2018, which is the latest data of the national survey. According to the definition in the survey studies, early adolescents are youth aged from 10 to 15 years old. Thus, we focused on early adolescents who were in an educational stage during the survey time of the year 2018.

### Measurements

#### Depressive symptoms

CFPS-2018 adopted a special 8-item questionnaire based on the standard Center for Epidemiological Studies Depression Scale (CES-D) ^12^ to screen depressive traits. This original questionnaire asked participants to report their emotional state in last week from 1 to 4, where “1” referred to “little or no (<1 day),” “2” referred to “not too many (1-2 days),” “3” referred to “sometimes half the time (3-4 days)” and “4” referred to “most of the time (5-7 days).” It contained one physiological symptom, two symptoms of anhedonia, and five symptoms of negative affection-cognition, consistent with CES-D’s design ^13^. Based on our previous study, this scale can be divided into two factors representing negative affection-cognition and anhedonia, by adding one physiological-aspect item to negative affection-cognition-related items, which is consistent with the structure of CES-D ^14^. The structural validity was stable across residential types (rural resident vs urban resident) and gender (male vs female) among early Chinese adolescents ^15^. Cronbach’s alpha was 0.71, indicating the acceptable internal consistency.

#### Negative experiences in school

CFPS program designed a scale containing 14 items to measure adolescents’ negative experiences in school on a Likert scale from 1 (“it does not describe me at all”) to 5 (“it describes me”). Seven items assess internal problems (e.g., “I have difficulties in concentrating myself”), and seven items evaluate external problems (e.g., “I quarrel with my peers frequently”). The reliability of the scale was acceptable (Cronbach’s alpha=0.79).

### Data Analysis

#### Descriptive statistics

The demographic statistical information would be depicted (including gender, age). And we would demonstrate the descriptive statistics for each measurement.

#### Node selection

We were interested in the relationship between early adolescents’ depressive symptoms and their negative experiences in school. However, previous studies only pointed out the psychometric characteristics for each variable measurement based on factor analysis. These latent variable analyses failed to provide a complex perspective to investigate the relationships between each concrete symptom or experience; instead, they only roughly measured general latent variables and the relationships between the common underlying variables. Network analysis has the advantage of delving deeper into the relationships among observed indicators. Therefore, first of all, we selected suitable symptoms and specific experiences in school as nodes for further network analysis. We adopted the “goldbricker” procedure in the R package of “network tools” to investigate the problematic topological overlap between variables. If two items are topologically connected with the other items without differential associations, it means these two items are the same, and one of them can be removed, or it is plausible to combine those two via principal component analysis (PCA) ^16^.

#### Within cluster centrality

The network outputted based on the Graphical Gaussian Model presents the relationships between nodes. An edge in the network describes partial correlations between two nodes after controlling for all the other correlations with left nodes in the network ^17^. Thus, the network was built by the “LASSO” graphic approach (gamma=0.05) in the R package bootnet ^18–20^. In this study, we computed the expected influence and strength centrality for nodes. Unlike strength centrality, which is the sum of the absolute values of the edge weights extending from a node to all others, expected influence keeps the value both positive and negative. In the psychopathology network, the positive or negative relationship is meaningful. We calculated the centrality for two clusters (depressive symptoms cluster and the cluster of negative experiences in school).

In addition, we computed the correlation-stability (CS) based on the case-dropping bootstrap method. CS describes the maximum portion of dropped data when the estimated centrality can still be highly correlated (r>0.7) with the original network. CS below 0.25 indicates unacceptable stability. If CS is above 0.5, the network’s stability is good.

#### Across-cluster (bridge) centrality

No network analysis of the relationship between children’s depressive symptoms and negative school experiences has been done before. In this case, bridge expected influence would be computed to identify negative school experiences linked to depressive symptoms.

## RESULTS

### Descriptive statistics

The average age of participants was 11.01 (SD=1.22), and the sample included 52.7% of males and 47.3% of females. The mean scores of depressive symptoms and negative school experiences were 1.48 (SD=0.41) and 2.27 (SD=0.61), respectively.

### Node selection

All the observations were standardized before entered into the analysis. The goldbricker function in R suggested among early adolescents’ depressive symptoms, the two anhedonia symptoms manifested the topological overlap. We then used PCA to combine these two symptoms. The goldbricker output did not suggest a reduction regarding adolescents’ negative experiences in school.

### Within cluster centrality

Within the cluster of depressive symptoms, the expected influence centrality summary revealed that item 7 (“I felt sad”), item 5 (“I felt lonely”), item 1 (“I felt depressed”), and item 2 (“I felt that everything I did was an effort”) were central items in screening depressive symptoms (see Figure 1). The CS coefficient was good (CS=0.67), which suggested the good stability of the network structure.

**Figure 1.**
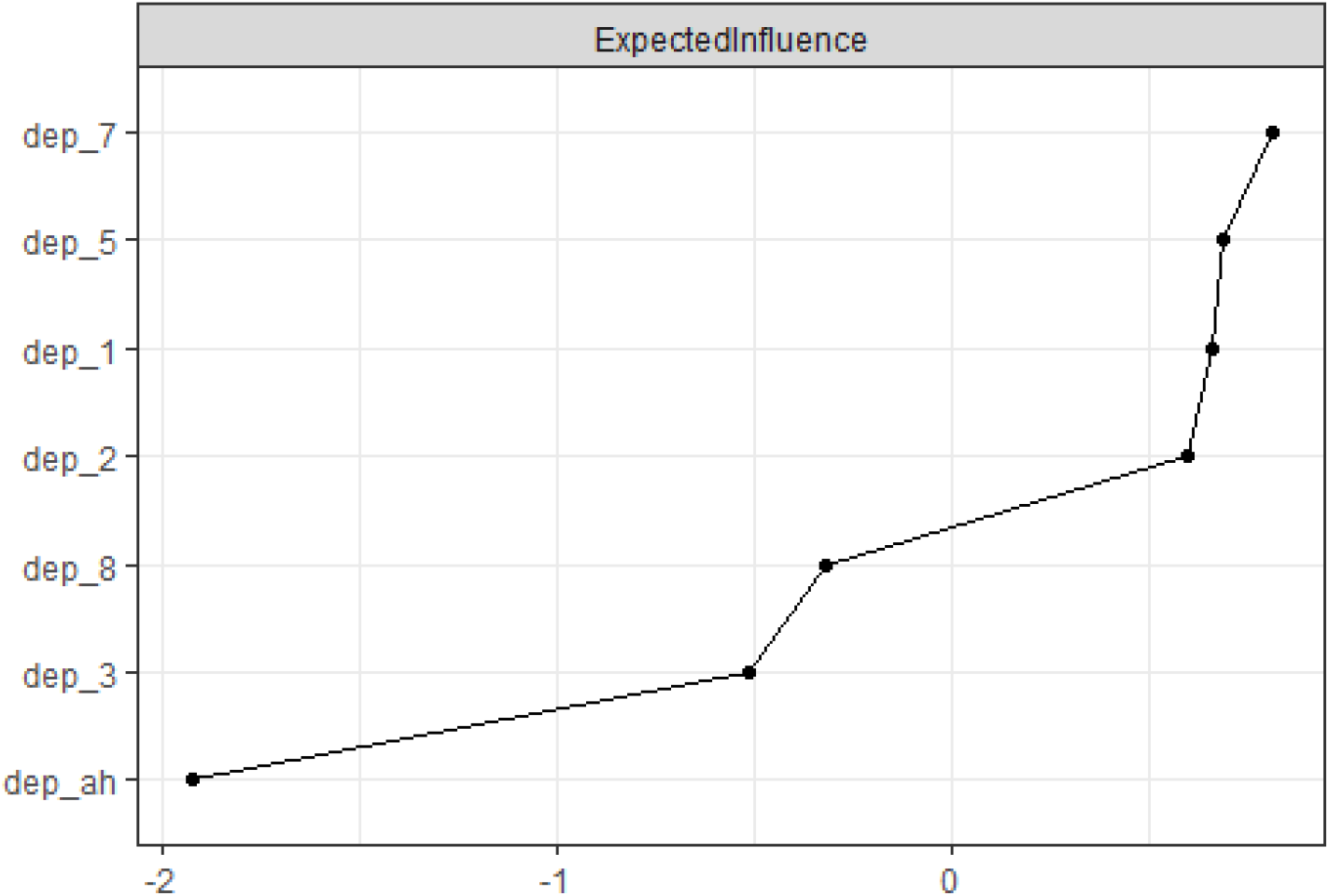
The illustration of the centrality (expected influence) of each depressive symptom in the depression-network

Within the negative school experiences cluster (Figure 2), central negative experiences in school were “I am afraid of examinations” (item 3), “ I have difficulties concentrating” (item 4), “I am in trouble because of blab” (item 12), and “I am worried about having no peers to play with at school”(item 11). The network was stable (CS=0.67).

**Figure 2.**
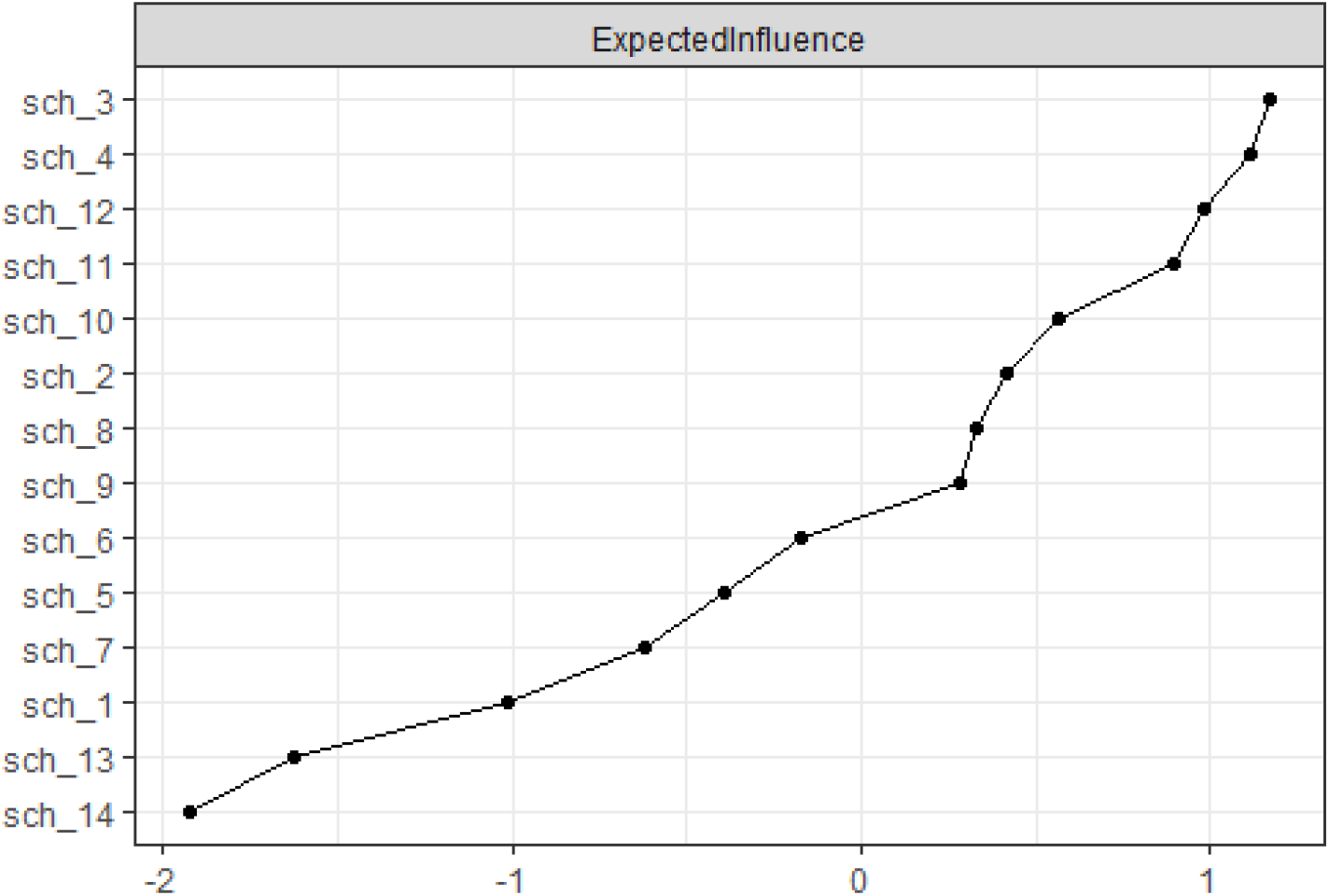
the illustration of the EI centrality of each negative experience in the school-experience-network

### Across-cluster (bridge) centrality

Across-clusters centrality suggested that the feeling of sadness in school was the most important node connecting to the cluster of adolescents’ depression. Difficulties in peer relationships including “I am worried about having no peers to play with at school” and “I am in trouble because of blab” were the next important nodes extending to depression.

## DISCUSSION

The current study using network analysis provides a better understanding of the relationship between adolescents’ depressive symptoms and their negative experiences in schools. This research first identified the most central depressive symptoms among Chinese adolescents and the central nodes of negative experiences in schools.

Within the cluster of adolescents’ depression, the most centralized nodes were subjective negative affections. The following central node was a negative cognition (“I felt that everything I did was an effort”). Anhedonia was the last symptom, furthest from the centre.

Within the cluster of “negative experiences in the school,” the most central nodes were the poor academic related performance (“I am afraid of examinations,” and “ I have difficulties in concentration”) and peer relationship problems (“I am in trouble because of blab,” and “I am worried about having no peers to play with at school”). These results comprehensively estimated Chinese adolescents’ negative experiences in schools, which have not been investigated before.

This study suggests a strong link between the negative emotional experience in school and depression. The findings demonstrate that the children’s lasting negative emotion and poor quality of peer relationships in schools are important indicators of depression-related mental health risk, which echo with numerous previous studies. School psychologists argue that schools provide a vital environment for children’s socialization process through competition and cooperation with peers, and the encouragements and supports from school decreases children’s anxiety and improves the level of self-esteem ^21^. Notably, a review work previously indicated depression and anxiety usually ruminate, which means supports from schools potentially reduce the risk of depression ^22^. Also, it is known that low self-esteem is associated with depression, and a longitudinal meta-analysis supported their cause-effect relationship, not simply correlation ^23^. Thus theoretically, social support provided by schools strongly and negatively correlates with adolescents’ depressive symptoms. Ample empirical evidence has verified this theory. For instance, poor quality of peer interpersonal interaction among school-aged adolescents has been found to increase the risk of depression ^24,25^. Furthermore, peer-reported aggressive behaviours in school have been associated with depression ^25^. Network analysis revealed that the lack of social support, especially the support from friends, correlates with adolescents’ internalizing symptoms, including depression ^16^.

### Limitations

However, due to the limitations of the open databases of CFPS, this study could not probe the relationship between negative experiences in school and the in-depth internalizing disorder. Further studies could combine anxiety and depression to conduct network analysis integrating internalizing problems, as such research is lacking in China.

### Conclusions

In conclusion, we found that negative affections and a negative cognition of “ I felt that everything I did was an effort” are the cores of Chinese early adolescents’ depressive symptoms. Anhedonia is the least central symptom. About their negative experiences in school, poor academic related performance (“I am afraid of examinations,” and “ I have difficulties in concentration”) and difficulties in peer relationships (“I am in trouble because of blab,” and “I am worried about having no peers to play with at school”) are central problems. Moreover, the negative emotional experiences in school and difficulties in peer relationships of early adolescents are the strongest predictors for their depression.

## IMPLICATIONS FOR SCHOOL HEALTH

This study has meaningful implications. The results offer a practical perspective to schoolteachers on school-aged adolescents’ mental health state, implying that teachers should pay attention to children’s negative affection and difficulties in peer relationships in school, as they are the most significant predictors towards depression.

### Human Subjects Approve Statement

All data came from the public database. Ethics approval and consent to participate were not applicable in this study. However, the data collection work of CFPS had been already approved by the Ethics Committees of the Institution of Social Science Survey, Peking University.

## Data Availability

Anyone who is interested in the dataset can send an email to contact the first author (Yi HUANG).

